# Sleep disturbances as risk factors for neurodegeneration later in life

**DOI:** 10.1101/2023.11.08.23298037

**Authors:** Emily Simmonds, Kristin S Levine, Jun Han, Hirotaka Iwaki, Mathew J Koretsky, Nicole Kuznetsov, Faraz Faghri, Caroline Warly Solsberg, Artur Schuh, Lietsel Jones, Sara Bandres-Ciga, Cornelis Blauwendraat, Andrew Singleton, Valentina Escott-Price, Hampton L Leonard, Mike A Nalls

**Author notes:** Ms Simmonds and Ms Levine contributed equally to this paper. Dr Escott-Price [ ] and Ms Leonard [ ] are joint corresponding authors.

## Abstract

The relationship between sleep disorders and neurodegeneration is complex and multi-faceted. Using over one million electronic health records (EHRs) from Wales, UK, and Finland, we mined biobank data to identify the relationships between sleep disorders and the subsequent manifestation of neurodegenerative diseases (NDDs) later in life. We then examined how these sleep disorders’ severity impacts neurodegeneration risk. Additionally, we investigated how sleep attributed risk may compensate for the lack of genetic risk factors (i.e. a lower polygenic risk score) in NDD manifestation.

We found that sleep disorders such as sleep apnea were associated with the risk of Alzheimer’s disease (AD), amyotrophic lateral sclerosis, dementia, Parkinson’s disease (PD), and vascular dementia in three national scale biobanks, with hazard ratios (HRs) ranging from 1.31 for PD to 5.11 for dementia. These sleep disorders imparted significant risk up to 15 years before the onset of an NDD. Cumulative number of sleep disorders in the EHRs were associated with a higher risk of neurodegeneration for dementia and vascular dementia. Sleep related risk factors were independent of genetic risk for Alzheimer’s and Parkinson’s, potentially compensating for low genetic risk in overall disease etiology. There is a significant multiplicative interaction regarding the combined risk of sleep disorders and Parkinson’s disease.

Poor sleep hygiene and sleep apnea are relatively modifiable risk factors with several treatment options, including CPAP and surgery, that could potentially reduce the risk of neurodegeneration. This is particularly interesting in how sleep related risk factors are significantly and independently enriched in manifesting NDD patients with low levels of genetic risk factors for these diseases.

**HIGHLIGHTS:**

- Sleep disorders, particularly sleep apnea, are associated with the risk of Alzheimer’s disease, amyotrophic lateral sclerosis, dementia, Parkinson’s disease, and vascular dementia in national scale biobanks.
- These sleep disorders imparted significant risk up to 15 years before the onset of a neurodegenerative disease.
- The cumulative number of sleep disorders in the electronic health records were associated with a higher risk of neurodegeneration related to dementia and vascular dementia.
- Sleep related risk factors are independent of genetic risk for Alzheimer’s and Parkinson’s, potentially compensating for low genetic risk in overall disease etiology.
- Significant multiplicative interaction exists regarding the combined risk of sleep disorders and Parkinson’s disease.

## INTRODUCTION

The World Health Organization has recognized sleep as a critical health state and health-related behavior [Accessed 30 Aug 2023] ^1^. One-fourth of Europeans have insomnia ^2^. Short-term daytime cognitive impairment is common for people with sleep disturbances that prevent them from getting adequate rest; individuals who experience sleep problems have been shown to have a greater risk of developing dementia ^3,4^. Obstructive sleep apnea (OSA), a primary sleep disorder, is a condition marked by nighttime airway collapse leading to brief lapses in breathing. Having OSA increases one’s risk of developing dementia in general and is particularly common with Alzheimer’s disease, occurring in about 50% of patients ^5^. Patients with insomnia also have a higher risk of dementia ^6^. Some evidence exists that sleep–wake and circadian disruption can occur early in the course of the disease or even precede the development of cognitive problems ^7^.

Although an association between sleep and neurological disorders is widely acknowledged, it is largely unclear how dysfunctions in sleep and circadian rhythms contribute to the etiology of neurodegeneration or whether they are a contributing cause or consequence of these conditions. Many individuals who develop dementia experience sleep problems following dementia onset, and there is evidence that these conditions are involved in a complex self-reinforcing bidirectional relationship (e.g., relationship between amyloid-β plaque accumulation and poor sleep) ^8^. As dysregulation of the circadian clock already occurs during the asymptomatic stage of the disease and could promote neurodegeneration, restoration of sleep and circadian rhythms in preclinical neurodegenerative disorders may represent an opportunity for early intervention to slow the disease course.

Sleep disturbances have also been associated with future risk of both cognitive impairment and AD pathology, and can be an early indication of pre-symptomatic stages of AD ^9,10^. Various types of dementia are associated with different types of sleep and circadian disturbances ^9–11^. Over 40% of AD patients are affected with a sleep disorder, and the prevalence and severity of sleep disorders increase with dementia severity ^12^. Sleep disturbance occurs very early in AD; even the preclinical stage of AD prior to cognitive symptoms is associated with worse sleep quality and shorter sleep duration ^13^. There is a similar prevalence of sleep disorders in frontotemporal dementia (FTD), but the pattern of rest/activity disturbance in AD and FTD differs ^14^.

PD and sleep are connected in complex ways that are not completely understood. Sleep-related symptoms may be one of the earliest signs of Parkinson’s disease ^15^. The majority of patients with rapid-eye movement sleep behavior disorder (RBD) eventually develop PD or another neurological condition^16^. A recent study found risk loci for RBD near known PD genes, such as SNCA and GBA^17^.

Lewy Body Disease (LBD) has the highest prevalence of sleep and circadian disturbances of any dementia, affecting approximately 90% of patients, with insomnia being the most prevalent ^18,19^. Sleep problems have also been reported in patients with ALS, including RBD and sleep apnea ^20^. Fatigue is a common symptom in patients with multiple sclerosis (MS), but sleep disorders are often overlooked in this population^21^.

To unravel the causal structure that relates to NDDs and sleep disorders, it is helpful to be able to examine individuals over as long a time period as possible, as well as differentiate between individuals who were diagnosed with sleep disorders pre- and post-NDD diagnosis. For this, medical health records are an invaluable tool, as they provide time-stamped information on all medical events, including symptoms, diagnoses, medication usage, and clinical interventions across an individual’s full time while enrolled in that system. Here, we use three national scale biobanks with massive sample sizes to disentangle the complexities of sleep and neurodegeneration while also examining the complex interplay between sleep, neurodegeneration, and genetics as part of this data mining effort.

## METHODS IN BRIEF

This study uses data from three biobanks: the Secure Anonymised Information Linkage (SAIL) databank^22^, as well as the UK Biobank (UKB)^23^ and FinnGen datasets^24^. SAIL is a repository of medical health records from Wales, UK, covering approximately 80% of all individuals living in Wales between 1970 and 2019 (Application Number 0998). UKB data were applied for and accessed via the UKB research and analysis portal (Application Number 33601), which hosts genotyping data from nearly 500,000 individuals from the UK. For controls in these cohorts, we used a subset of age-matched (baseline age greater than 45 years) unrelated individuals of European ancestry who did not have an NDD diagnosis or a family history of NDDs. Summary statistics from FinnGen, a nationwide Finnish biobank with genotyping data available for over 377,000 individuals, including hazard ratios for sleep disorder endpoints, were downloaded through FinnGen’s Ristey’s portal in May 2023, to further replicate our results.

We used the first reported date of an ICD10 sleep code (F51 or G47) as a sleep disorder exposure, excluding any ICD10 codes occurring after an NDD diagnosis. Any NDD/sleep disorder association with less than five pairs (i.e. individuals with a sleep disorder diagnosis who also later developed an NDD) were excluded to reduce low statistical power analyses.

All statistical analyses, including logistic regressions, cox proportional hazard models, and polygenic risk score associations were adjusted for age, sex, and socioeconomic status when possible. All reported p-values were adjusted using false discovery rate correction.

A summary of the study design and analyses can be found in **Figure 1**. See **Table 1** for case numbers and sex breakdowns for each of the six NDDs and **Supplementary Table 1** for information about the endpoints and sleep groupings analyzed in this study.

**Figure 1:**
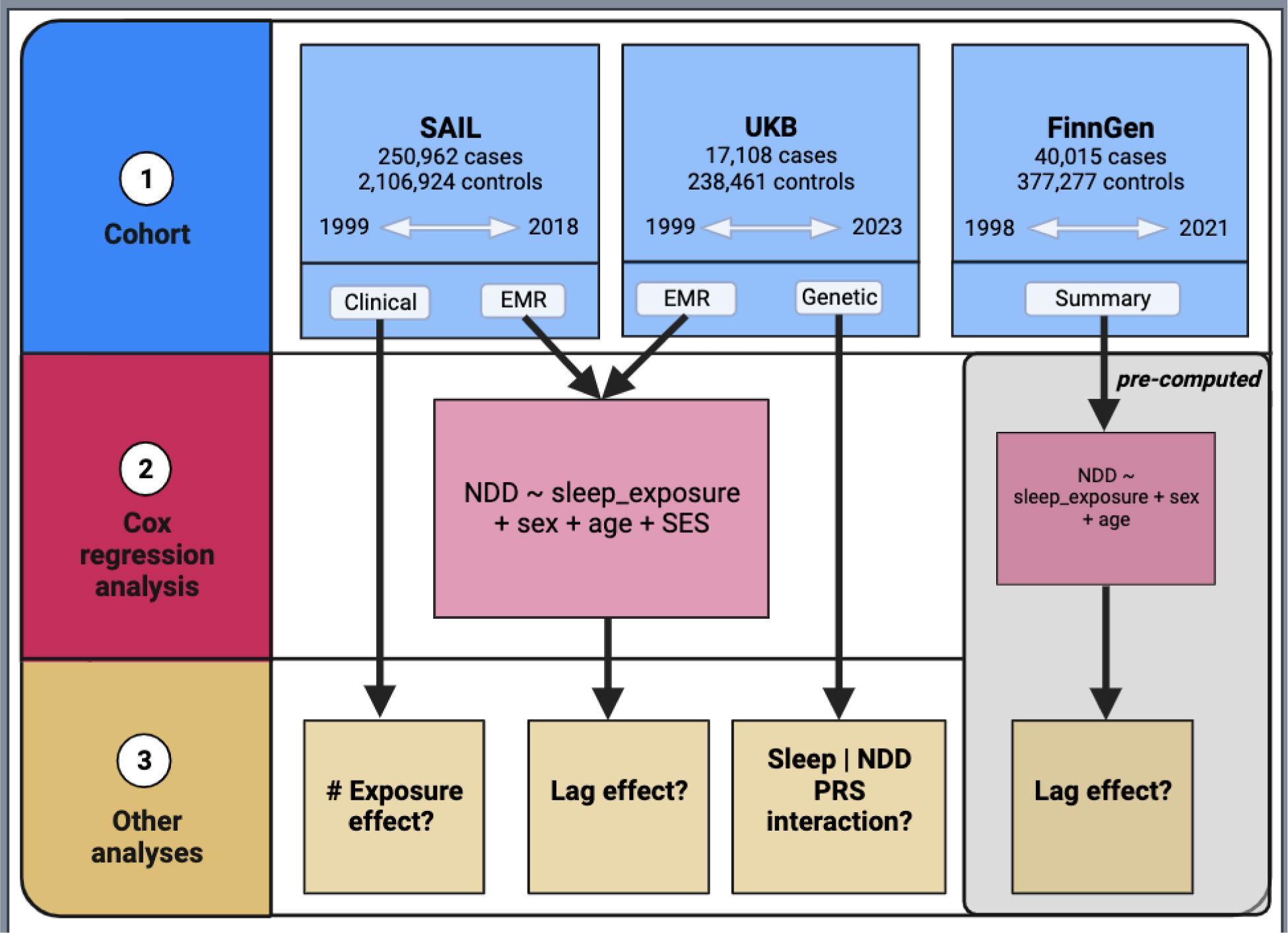
**Study Design** – A summary of the study design and analyses performed.

Please see the **STAR METHODS** section below for additional details as well as the **Key Resource Table** in that section for available data and code.

## RESULTS

### Prior sleep disorders are associated with late life neurodegeneration

After multiple test corrections, six pairs of sleep disorders and NDDs were shown to have significant associations in all three datasets, as summarized in **Table 2**. AD, dementia, and PD were all significantly associated with the code G47 which encompasses sleep disorders associated with circadian rhythm, such as narcolepsy, apnea, hyper and parasomnia, as well as cataplexy and movement related sleep issues. G47 coding associated with these diseases exhibited hazard ratios (HRs) ranging from 1.31 (95% CI 1.08-1.59 for PD in SAIL) to 2.80 (95% CI 1.76-4.47 for PD in FinnGen). For a graphical summary of these results, see **Figure 2**.

**Figure 2:**
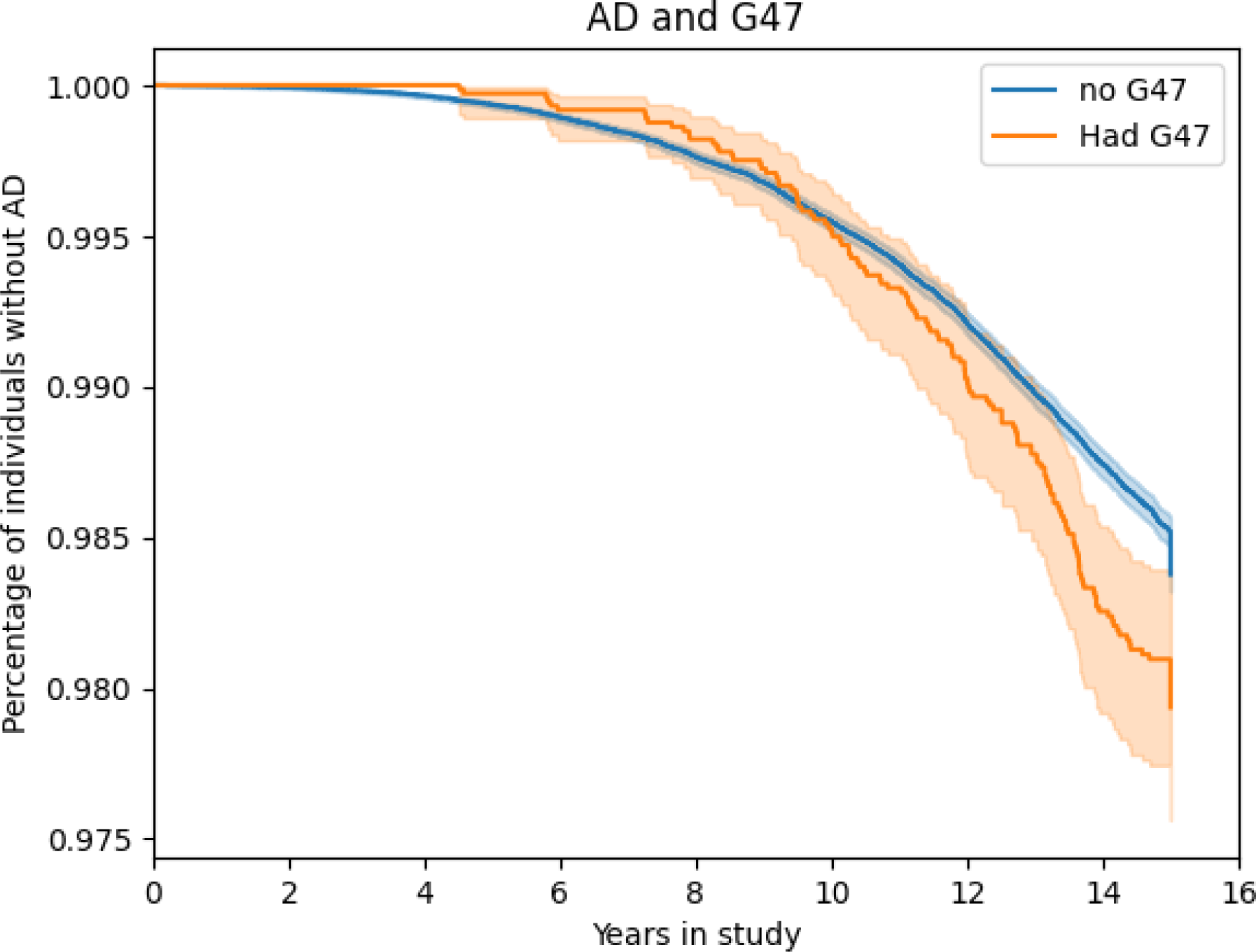
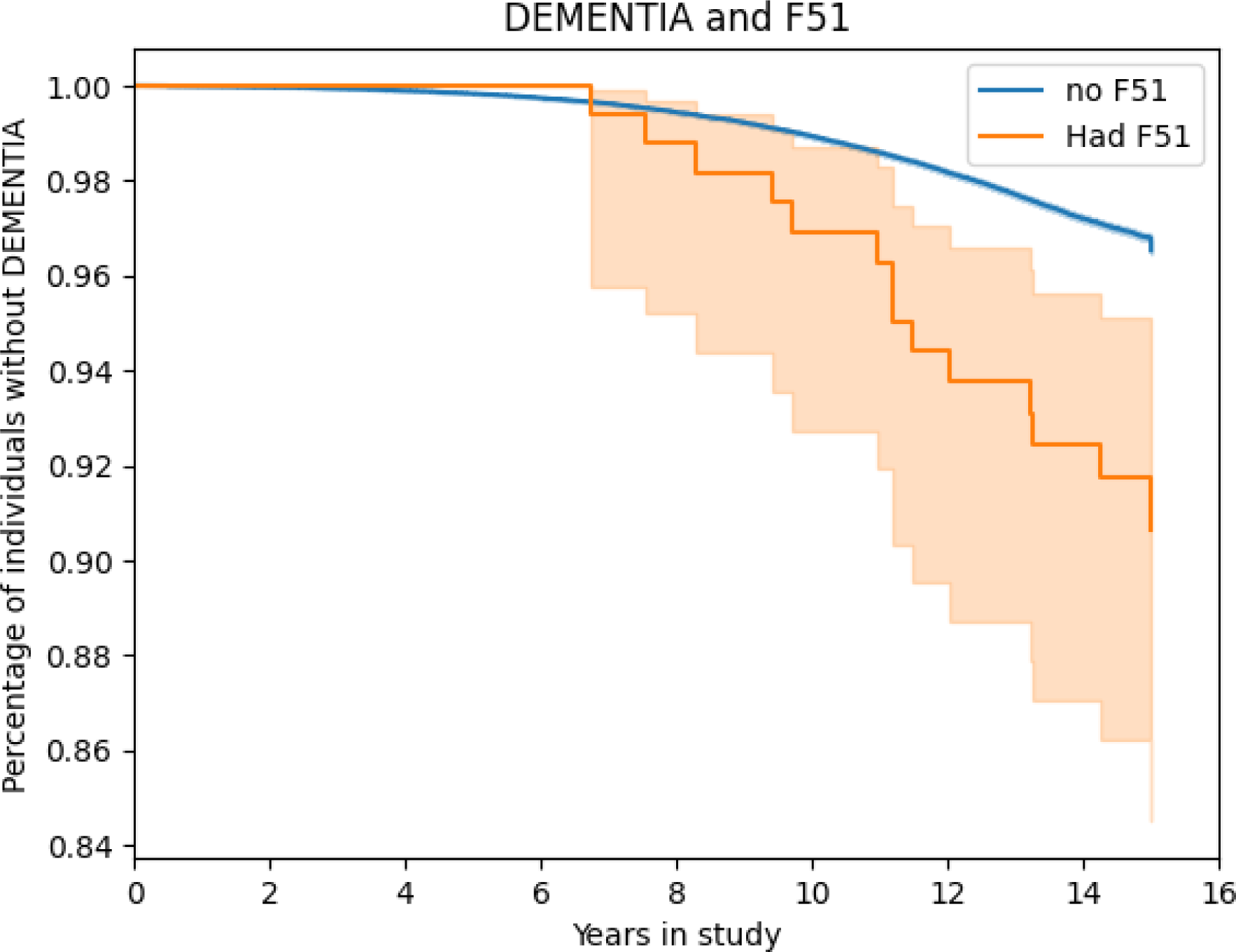
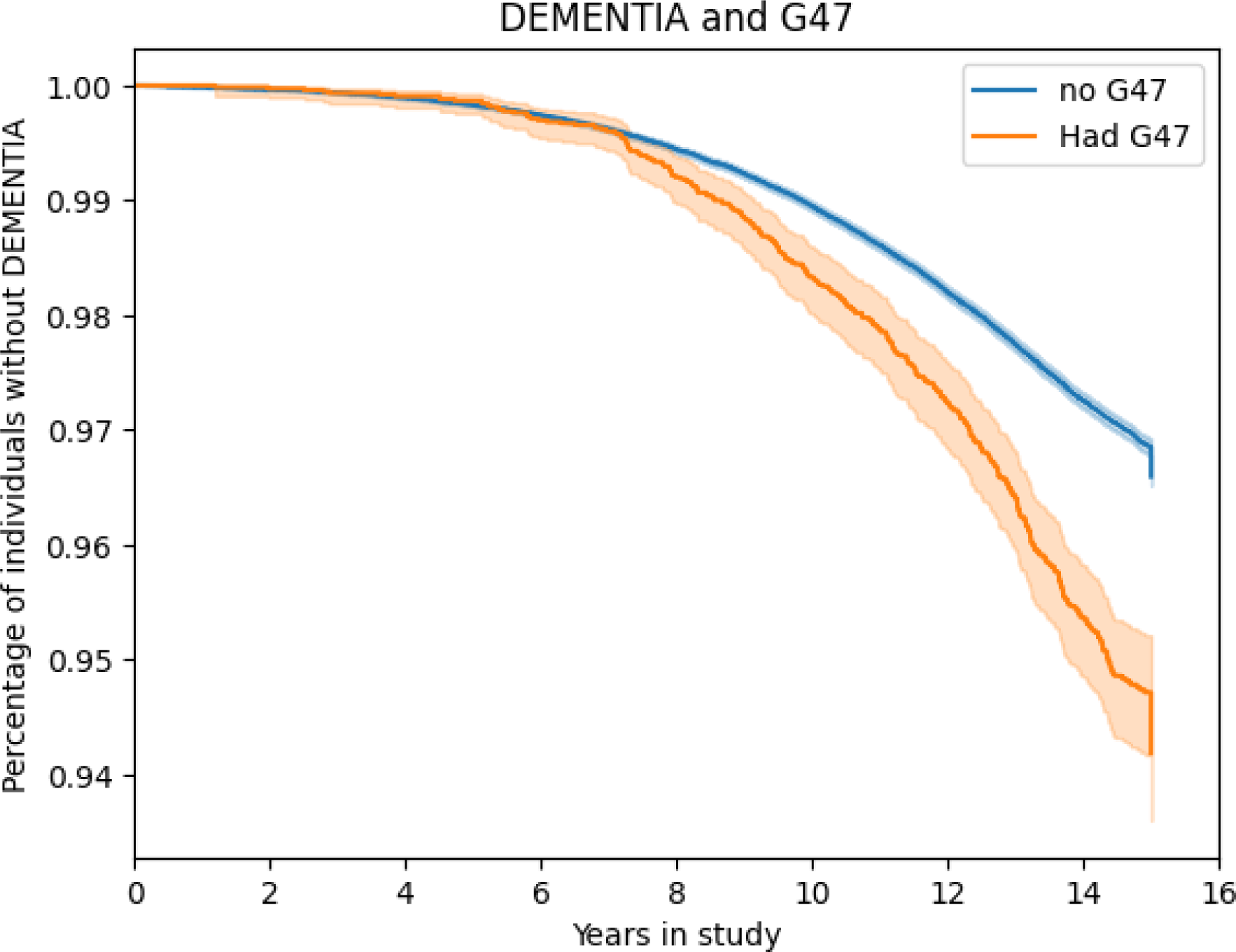
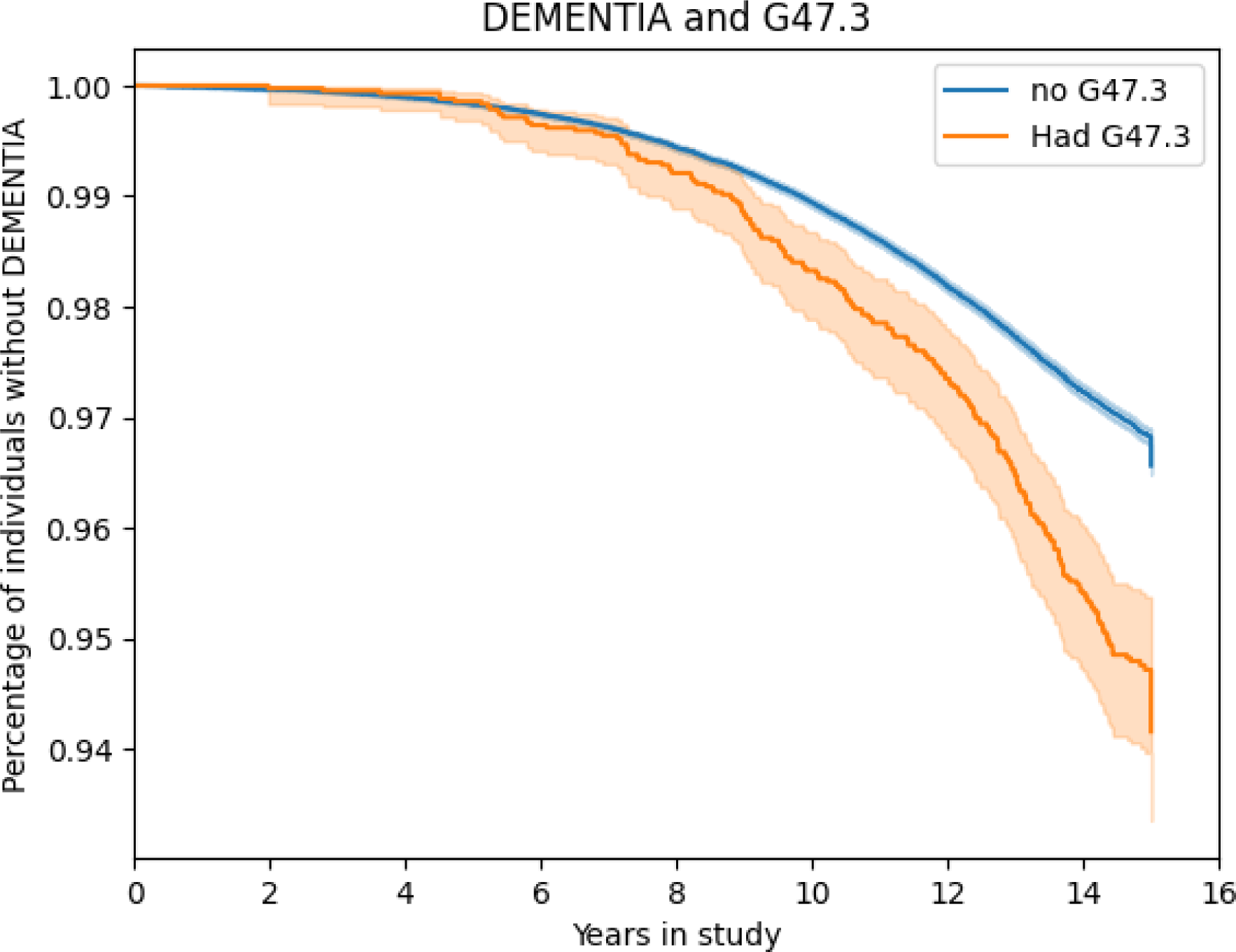
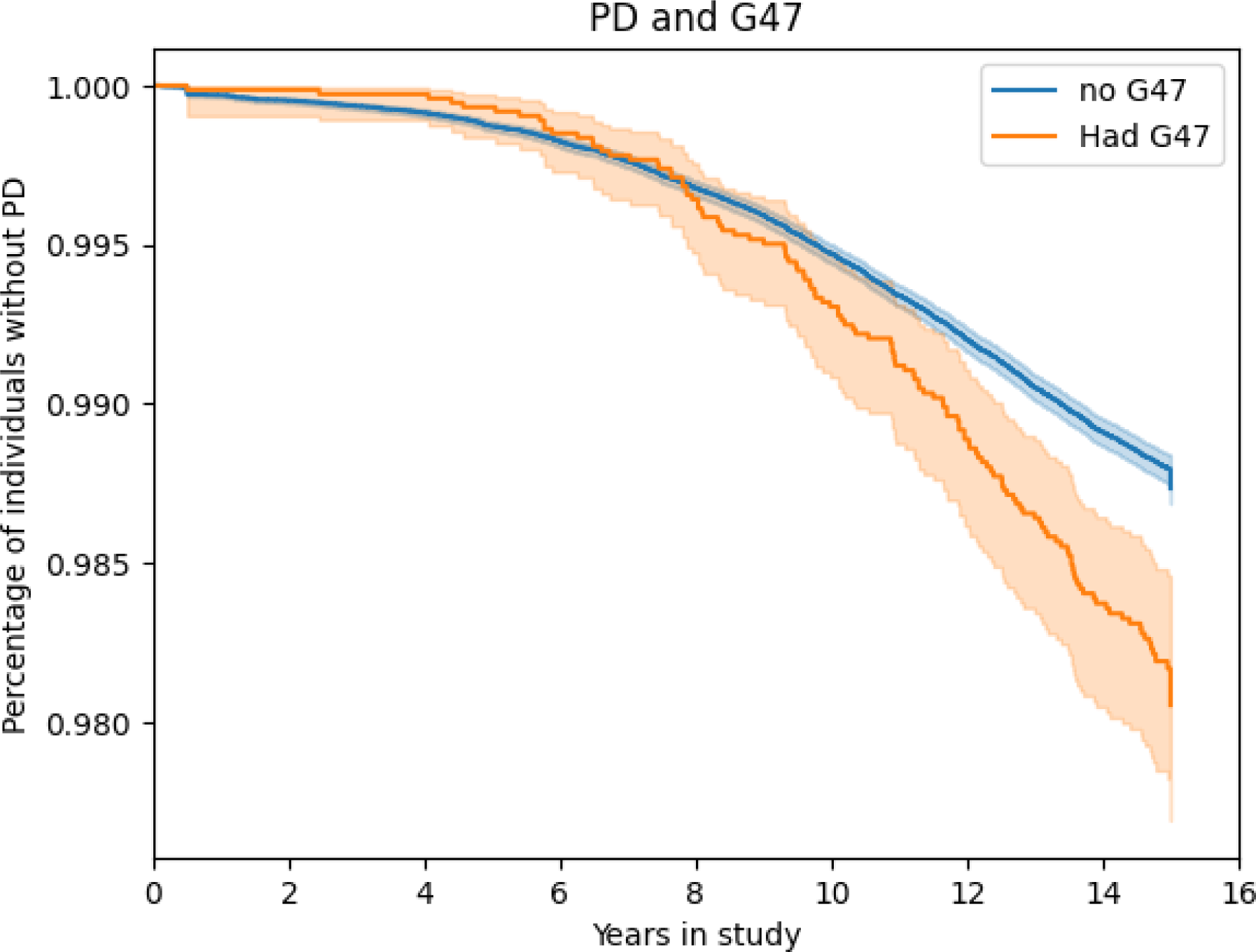
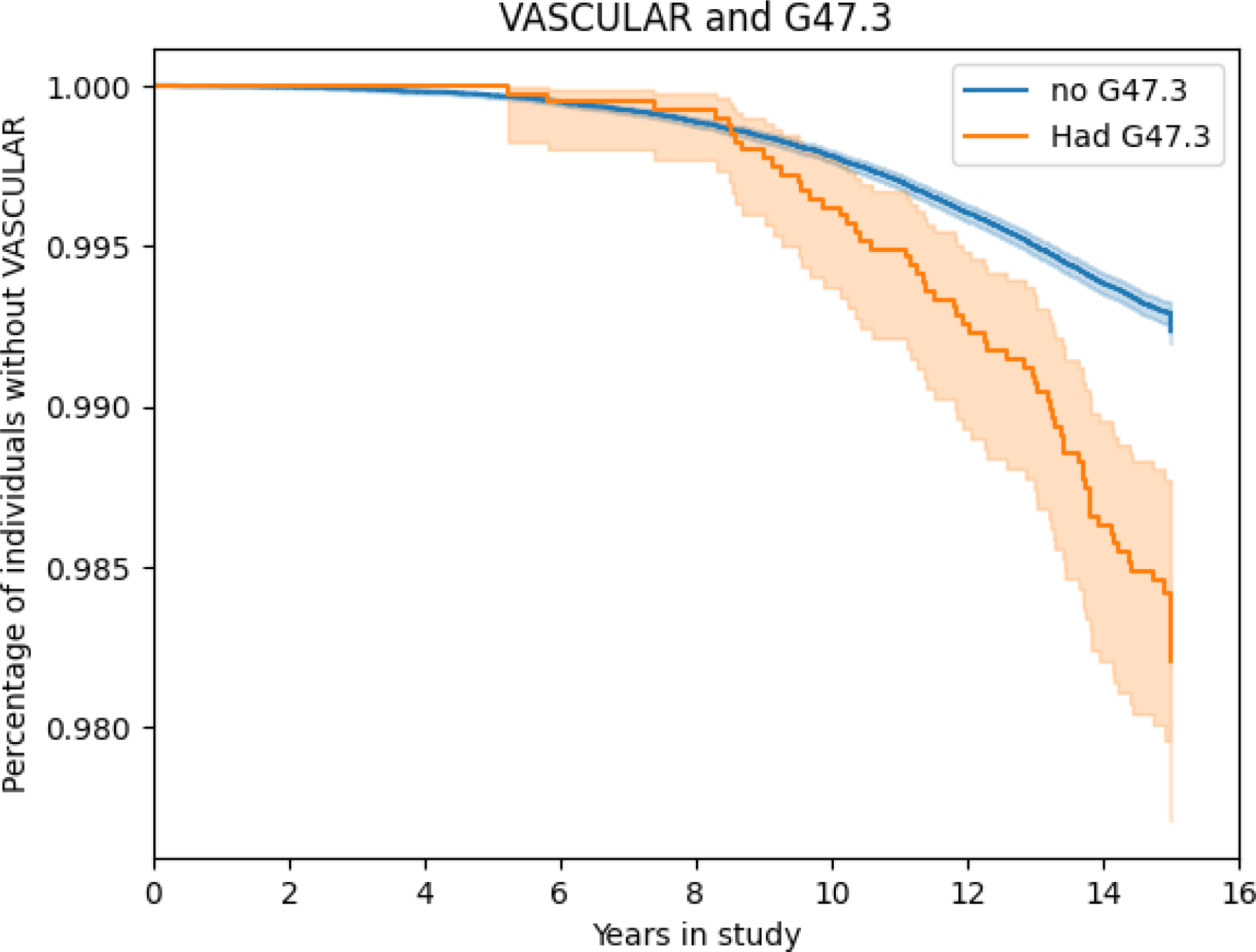
Kaplan-Meier Plots for Replicated NDD/Sleep code pairs. Kaplan-Meyer plots showing longitudinal differences in disease risk between individuals with sleep disturbances and those without. Only replicated pairings are plotted. **G47**: Sleep disorders associated with circadian rhythm, such as narcolepsy, apnea, hyper and parasomnia, as well as cataplexy and movement related sleep issues. **F51**: Non-organic sleep disorders, such as nightmares and generalized insomnia (without substance abuse) **G47.3**: Sleep apnea

Non-organic sleep disorders under ICD10 code F51 include mental, behavioral, and neurodevelopmental conditions such as nightmares and generalized insomnia (without substance abuse). They are significantly associated with increased risk of dementia across all three datasets. These associations ranged in HRs from 2.61 (95% CI 1.55-4.41 in UKB) to 5.11 (95% CI 2.75-9.51 in SAIL). In general, these are the largest HRs observed in this report.

Sleep apnea (ICD-10 code G47.3) specifically was associated with dementia and vascular dementia, suggesting that there could be cardiometabolic connections to risk. HRs ranged from 1.67 (95% CI 1.46-1.92 for general dementia in UKB) to 2.45 (95% CI 1.84-3.26 for vascular dementia in FinnGen).

Eight pairs of sleep disorders and NDDs were shown to have significant associations in two datasets, also summarized in **Table 2**. Sleep apnea was significant for ALS and PD, with HRs ranging from 1.42 for PD in UKB (95% CI 1.13-1.79) to 2.66 in for ALS in FinnGen (95% CI 1.93-3.67). These associations for ALS should be taken with caution as ALS is relatively rare across biobanks; no ALS data was available in SAIL. Non-organic sleep disorders (F51) were associated with PD and vascular dementia, with HRs ranging from 2.52 (95% CI 1.34-4.76 for PD in FinnGen) to 5.99 (95% CI 2.85-12.58 for vascular dementia in UKB). For PD, G47.8 and G47.9 were also significantly associated in two cohorts, with HRs ranging from 2.06 in SAIL to 8.07 in UKB (95% CI 1.40-3.04 and CI 4.03-16.17 respectively). G47.8 and G47.9 are both “other” sleep disorders codes. Since REM sleep disorder was not given an ICD10 code until 2016, it is possible that some of these codings may reflect that disorder.

While not significant in UKB or FinnGen, the protective association between G47 EHR codes and MS was highly significant in SAIL at a p-value of 9.18E-06 and HR 0.41 (95% CI 0.28-0.60), which may connect to reports of prodromal fatigue in MS.

### Risk of neurodegeneration persists up to 15 years before disease onset

In order to better assess the temporal effect on risk over fifteen years of follow-up, we split our cohorts into strata, dividing the cohorts by those who received a sleep disorder code in the EHR less than one year, one to five years, or five to fifteen years prior to NDD diagnosis. We then re-evaluated the risk of NDDs for the six significant pairings described above. These associations are detailed in **Table 3**.

The largest subset HRs were found less than one year before diagnosis, with HRs ranging from 1.73 for G47/PD in SAIL (95% CI 1.10-2.72) to 21.24 for F51/dementia in FinnGen (95% CI 9.32-48.37). In these cases, the sleep disorders may be early symptoms of neurological disease.

However, for AD, dementia, and PD, the G47 pairing remained significant 5-15 years before diagnosis, with HRs ranging from 1.44 (for AD 95% CI 1.13-1.83 in UKB) to 2.54 (for AD 95% CI 1.59-4.08 in FinnGen). We then broke the groupings down further for UKB and SAIL, looking at 5-10 and 10-15 year intervals. For AD and dementia, the association remained significant 5-10 years before diagnosis in both cohorts. For dementia, the G47 association also remained significant 10-15 years before diagnosis in UKB, with an HR of 1.56 (95% CI 1.27-1.94). For PD, the G47 association remained significant 5-10 years before diagnosis in UKB and 10-15 years before diagnosis in SAIL, with an HR of 1.70 and 1.84 respectively (95% C I1.22-2.38 and CI 1.20-2.82). The G47.3 (sleep apnea) coding also remained significant for dementia and vascular dementia at 5-10 years before NDD diagnosis, with HRs ranging from 1.71 (dementia 95% CI 1.33-2.20 in UKB) to 3.25 (vascular dementia 95% CI 1.98-5.33 in SAIL).

### More severe sleep disturbances impart higher risk

In SAIL, data was available for G47 coded sleep disorders with more detail, so we used the number of times an individual received a sleep disorder ICD10 code in their EHR as a proxy for disease severity. We compared individuals risk estimates at one, two, and three or more recorded sleep disorder codes during follow-up. Those who experienced zero incidents of sleep disturbances were used as the reference group. In general, the trend for increasing risk with recurrent sleep disturbances codes was significant for dementia (HR 1.42, 95% CI 1.34-1.49, p-value 8.9E-36) and vascular dementia (HR 1.56, 95% CI 1.41-1.73, p-value 1.50E-17). This trend was mirrored for F51 coded sleep disorders in dementia as well (HR 2.19, 95% CI 1.51-3.18, p-value 1.01E-04). It is also worth noting a significant inverse relationship between G47 coded sleep disorders and MS (HR 0.59, 95% CI 0.47-0.75, p-value 4.19E-05), with more reported sleep disturbances associated with even less risk of MS, likely due to fatigue related symptoms. These association tests are detailed in **Table 4**.

### Sleep disturbances compensate for low genetic risk

We also evaluated potential genetic interactions between the PRS for PD and AD with sleep disorders. Distributions of PD PRS were significantly different between cases of PD with a sleep disorder versus cases that did not have either F51 (T-test: statistic −4.16, p-value 3.29E-05) or G47 (T-test: statistic −3.74, p-value 1.89E-04) sleep disorders. For AD, distributions of AD PRS (including APOE) were significantly different for G47 (T-test: statistic −2.38, p-value 1.75E-02). When we excluded APOE from the AD PRS, results were no longer significant (T-test: statistic −1.00, p-value 0.32). There were no significant differences in distribution for AD and F51. These results are graphically summarized in **Figure 3**.

**Figure 3:**
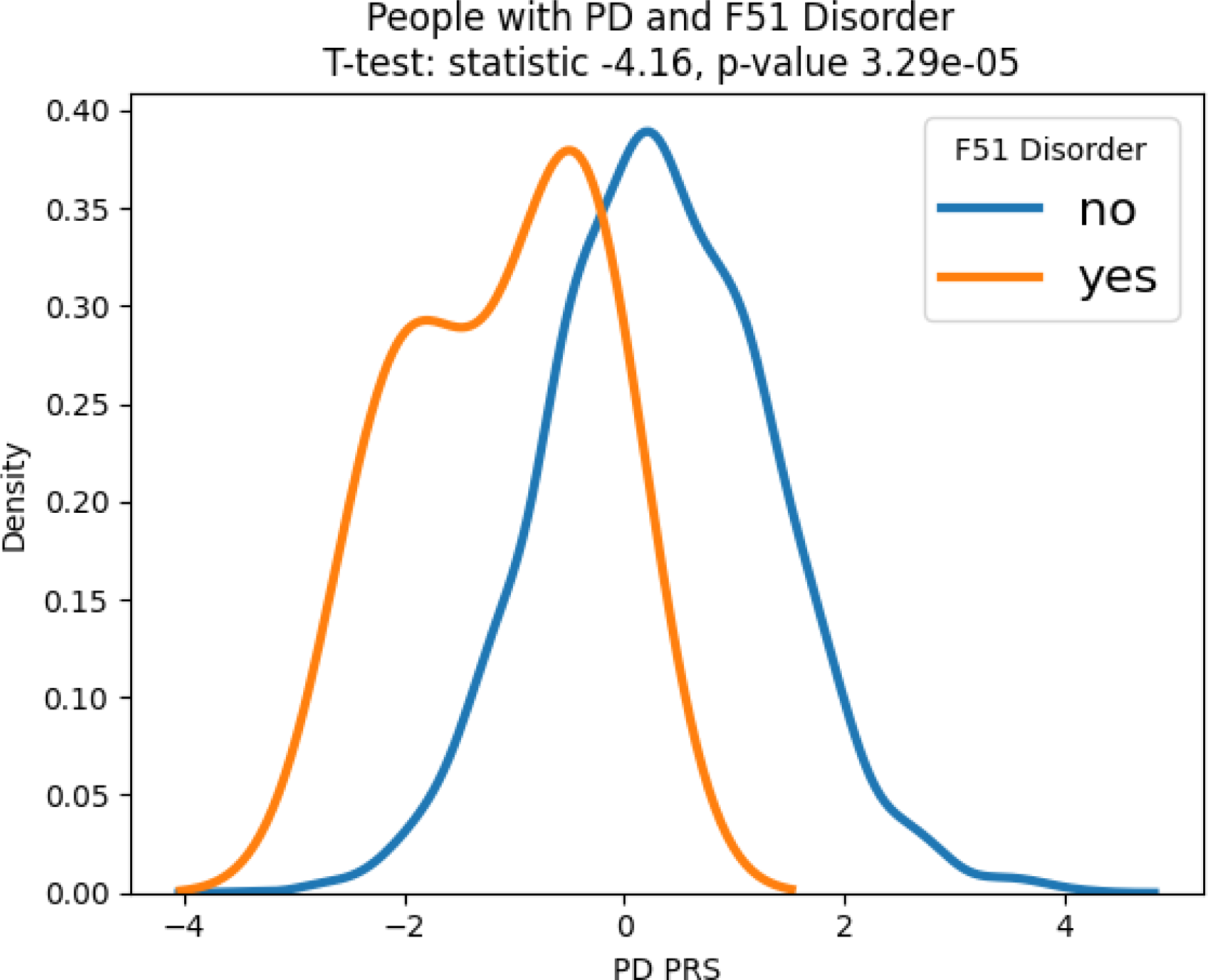
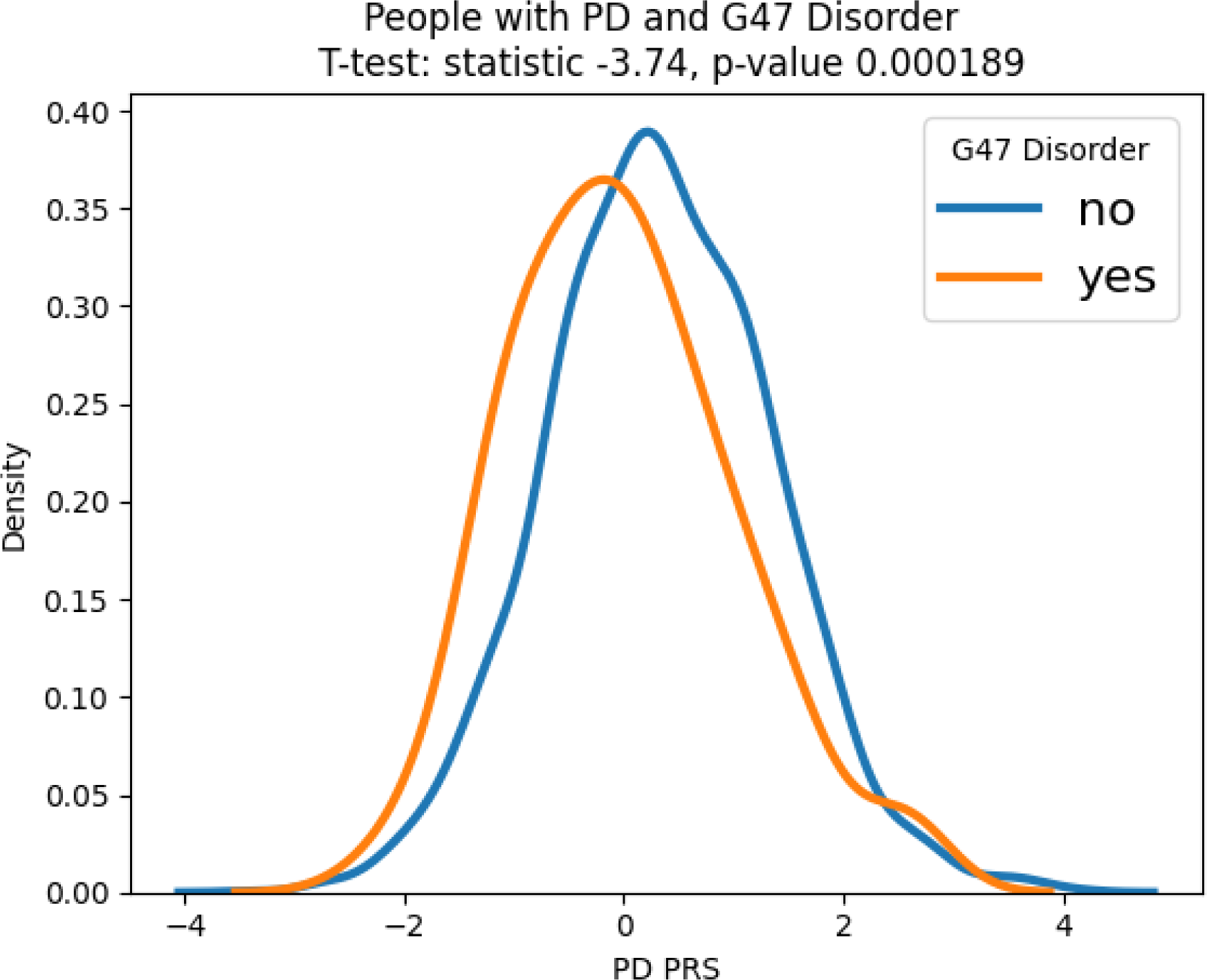
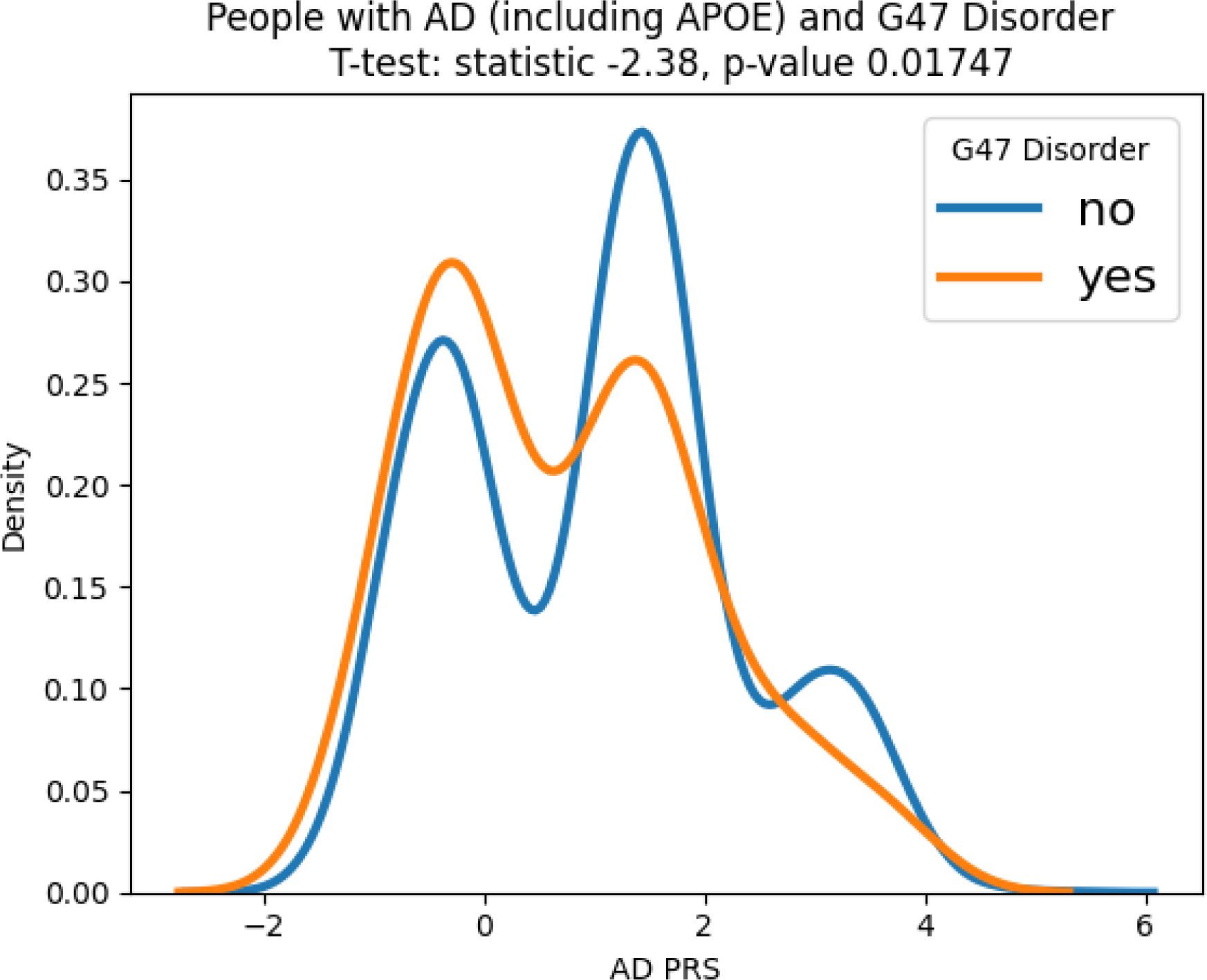
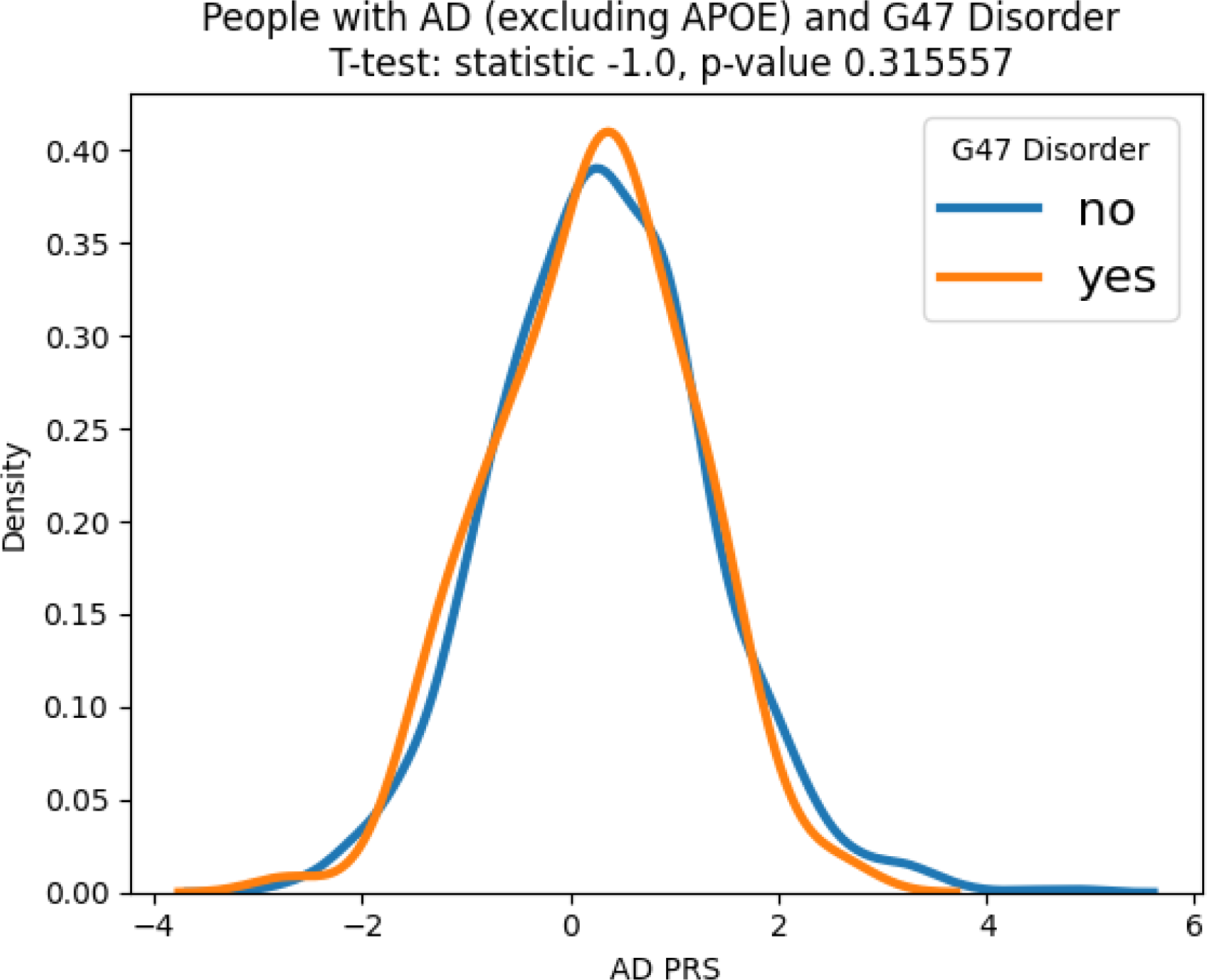
PRS density plots for NDD cases with and without a sleep disorder. We looked at PD PRS and AD PRS (with and without APOE) and found that PRS are differently distributed between cases of NDDs with and without sleep disturbances. These differences were significant for PD & F51, PD & G47, and AD (with APOE) & G47. AD PRS without APOE was not significant.

This led us to formally test for interactions between the PRS for each NDD and the sleep disorders, as detailed in **Table 5**. For each model, the sleep disorder was generally independent of the PRS as they remained significantly associated with the NDD when included in the model together, except for F51 codes and the PD PRS, which attenuated the F51 code parameter estimate’s p-value to 0.12. For both the F51 and G47 PD interaction models, the interaction term was significant and protective even after adjusting for the codes and PRS separately (ORs of 0.23 and 0.73, p-values of 5.50E-04 and 5.58E-04 respectively).

We also tested each of the interaction models with age at onset of either PD or AD as the outcome and no interaction terms were significant.

## DISCUSSION

In this work, we have gained insights into the complex relationship between sleep disturbances and late-life risk of neurodegeneration. While previous studies have investigated the associations between sleep and neurodegenerative diseases, this is the first large-scale survey that has shown replicated associations for multiple NDDs across multiple biobanks. This risk persisted for up to 10-15 years before NDD diagnosis for dementia, and 5-10 years for AD and PD. Severity of sleep disorders tended to increase risk as well. Finally, we looked into the interplay between sleep related risk and genetically derived risk, providing evidence that sleep disorders and genetics interact in PD to some degree, but are likely separate mechanisms leading to the same outcome in AD.

Modifiable risk factors such as maintaining proper sleep hygiene can potentially help reduce an individual’s risk of developing a neurodegenerative disease with a number of lifestyle, chemical, and mechanical interventions that are widely available. The circadian clock and sleep can influence a number of key processes involved in neurodegeneration, suggesting that these systems might be manipulated to promote healthy brain aging ^9^. In addition, there is some evidence that positive airway pressure therapy for sleep apnea is associated with a lowered risk of AD or dementia diagnosis.^25^

Medications used in neurodegenerative conditions can also affect sleep architecture. Dopaminergic medications used in PD can improve motor symptoms, but may lead to sleep disturbances, including insomnia and restless leg syndrome. Some antidepressant medications can lead to insomnia^26^. Cholinesterase inhibitors, such as donepezil, can improve cognitive and behavioral symptoms in people with Alzheimer’s but may have side effects like vivid dreams, nightmares, and insomnia ^27^. However, medications used to improve sleep may slow or stop the progression of neurodegeneration ^28^, suggesting this as an area for future research.

This report is not without its limitations. Summary data level access to FinnGen is excellent, although participant-level access could have facilitated additional modeling efforts. All of our datasets use diagnoses based on medical billing codes and not bloodwork or other assays. This coupled with data sparsity, can cause issues with some analyses. For example, the ICD10 code for REM sleep behavior disorder (RBD) is G47.52, which means it should be included in the G47 coding. However, while we were able to break out the specific coding for sleep apnea (G47.3), the specific code for RBD does not appear in UKB, so we were unable to analyze it separately.

When comparing the results of SAIL and UKB, the differences between the two datasets in the results may be due to available diagnosis codes and differences in sample makeup. In particular, incidence of dementia in the UKB may be different compared to the SAIL dataset because the UKB participants are volunteers who have not yet reached old age and are self-selected. We also did not account for the duration and for medication usage when considering severity, using only history and numbers of sleep disorders as a proxy for severity.

In addition, all the cohorts analyzed are predominantly European ancestry and may not be globally representative. Finally, we only had access to genetic data in one dataset on a participant level. Therefore, tests of interaction and independence of genetics and sleep related risk have yet to be formally replicated.

Sleep disorders were still mostly significantly associated with the tested NDD phenotype even after correction for PRS, suggesting that while there are some known common genetic risk factors for sleep disorders and disease (such as *GBA1* shared between RBD and PD)^29^, the risk observed between sleep disorders and NDDs does not appear to be due to underlying genetic risk factors alone. Not only does this report show a propensity for modifiable risk factors like sleep hygiene to help influence positive trajectories for late life brain health, but our analysis suggests interesting ramifications for precision medicine research. Since many trials focus on genetic and genomics supported therapeutics, sleep’s effect on disease etiology in potential trial recruits with low genetic risk burdens could be important as part of patient stratification models to avoid confounding and to increase efficiency ^30^.

## Supporting information

Table 1: Summary of participants in the study

Table 2: Replicated Sleep/NDD pairs

Table 3: Lags for replicated associations

Table 4: Multi-Exposure for G47 and F51 sleep disorders

PRS and Sleep interaction analysis

## Data Availability

This paper analyzes existing, publicly available data: the Secure Anonymised Information Linkage (SAIL) databank, as well as the UK Biobank (UKB) and FinnGen datasets.

https://saildatabank.com/

https://www.ukbiobank.ac.uk/

https://www.finngen.fi/en/access_results

## SUPPLEMENTAL INFORMATION

[Links](https://drive.google.com/drive/folders/1y1B32UFikMoGKFycpxFL9VD8adGcoQF4?usp=sharing) to google drive with additional tables.

## ACKNOWLEDGMENTS

This research was supported in part by the Intramural Research Program of the NIH, National Institute on Aging (NIA), National Institutes of Health, Department of Health and Human Services; project number ZO1 AG000535, as well as the National Institute of Neurological Disorders and Stroke. This work was also supported by the Dementia Research Institute [UKDRI supported by the Medical Research Council (UKDRI-3003), Alzheimer’s Research UK, and Alzheimer’s Society], Welsh Government, Joint Programming for Neurodegeneration (MRC: MR/T04604X/1), Dementia Platforms UK (MRC: MR/L023784/2) and MRC Centre for Neuropsychiatric Genetics and Genomics (MR/L010305/1). This research has been conducted using the UK Biobank Resource under application number 33601. We want to acknowledge the participants and investigators of the FinnGen study and thank them for their hard work and generosity. This study makes use of anonymised data held in the Secure Anonymised Information Linkage (SAIL) Databank. This work uses data provided by patients and collected by the NHS as part of their care and support. We would also like to acknowledge all data providers who make anonymised data available for research. We wish to acknowledge the collaborative partnership that enabled acquisition and access to the de-identified data, which led to this output. The collaboration was led by the Swansea University Health Data Research UK team under the direction of the Welsh Government Technical Advisory Cell (TAC) and includes the following groups and organizations: the SAIL Databank, Administrative Data Research (ADR) Wales, Digital Health and Care Wales (DHCW), Public Health Wales, NHS Shared Services Partnership (NWSSP) and the Welsh Ambulance Service Trust (WAST). All research conducted has been completed under the permission and approval of the SAIL independent Information Governance Review Panel (IGRP) project number 0998.

## AUTHOR CONTRIBUTIONS

E.S., K.S.L., H.L.L., V.E.P., M.A.N., and H.I. designed the study; E.S., K.S.L., H.L.L., C.B., V.E.P. and M.A.N. acquired and processed data; E.S., K.S.L., M.A.N., H.I., V.E.P. and H.L.L. analyzed data; all authors contributed to interpreting data and writing and editing of the manuscript.

## INCLUSION AND DIVERSITY

We support inclusive, diverse, and equitable conduct of research.

## TABLE LEGENDS

**Table 1**: Summary of participants in the study.

**Table 2**: Replicated Sleep/NDD pairs.

**Table 3**: Lags for replicated associations.

**Table 4**: Multi-Exposure for G47 and F51 sleep disorders.

**Table 5**: PRS and Sleep interaction analysis.

## STAR METHODS

### KEY RESOURCE TABLE

Please refer to the Key Resource Table.

### RESOURCE AVAILABILITY

#### Lead contact

Further information and requests should be directed to and will be fulfilled by the lead contact, Valentina Escott-Price, Hampton Leonard and Mike Nalls.

#### Materials Availability

FinnGen data is available in the link here [https://risteys.finngen.fi/] and was last accessed in July 2023, using Risteys R10. The UKB data is available here [https://www.ukbiobank.ac.uk/] and last accessed on August 30th 2023. Both the discovery and replication phase dataset are covered by the relevant national IRB groups.

#### Data and Code Availability

Notebooks containing code used in this analysis can be found in the GitHub link here: [https://github.com/NIH-CARD/NDD_x_sleep]. This paper analyzes existing, publicly available data. In addition, complete summary statistics describing these data/processed datasets derived from these data have been deposited in the supplementary materials connected to this publication and are publicly available as of the date of publication.

### METHODS DETAILS

Our initial study design included a discovery and replication phase, using time-aware data from three biobank scale datasets: SAIL, UKB, and FinnGen. Details of all codings used are given in Supplementary_Table_1.

#### SAIL Databank

The Secure Anonymised Information Linkage (SAIL) databank is a virtual platform that facilitates the linkage of various medical datasets using anonymised IDs. Diagnoses of disorders in SAIL came from the Patient Episode Database for Wales (PEDW), records from clinicians and hospital staff, and from the Welsh Longitudinal General Practitioner dataset (WLGP), records from primary care physicians of diagnoses, treatments, symptoms, and referrals. Demographic data on sex, age, address, and death were sourced from the Welsh Demographic Services Database (WDSD), and from WLGP.

To identify disorder diagnoses, PEDW was queried using ICD10 codes, and WLGP using NHS read codes (CVT2,3). We did not include neurodegenerative disorders from the outpatient data (OPDW) since only 0.1% of dementia cases are present in this data, and it only provides the attendance date which would not be a robust diagnosis date. Due to the lack of accurate diagnosis date, we also did not include dementia diagnoses from death data (ADDES).

The study period used was January 1^st^ 1999 to December 31^st^ 2018. Therefore, for inclusion, individuals needed to be alive at the start of 1999. Additionally, to ensure that all included individuals had a chance of developing dementia before the end of the study period, we decided to include only those who were aged at least 45 on January 1^st^ 1999. We also excluded any individuals missing age and sex information and individuals who did not have a Welsh address.

#### UK Biobank

The UK Biobank (UKB) is a large prospective cohort of individuals from the UK recruited between 2006-2010 ^41^. Diagnoses of disorders in UKB were derived using ICD10 codes as in SAIL. Information on sex and Townsend deprivation scores was available for all individuals. We left-censored any data occurring before January 1, 1999, to match SAIL’s records and right-censored any data occurring after NDD diagnosis. Any ICD10 codes with fewer than 5 cases were excluded from analysis.

Individuals from the UK Biobank were selected for this study if they self-reported as white British and were of similar genetic ancestry by principal component analysis (UK Biobank field 22006) and were unrelated (kinship coefficient < 0.0884). Genetic data in UKBB is already imputed using the haplotype reference consortium (HRC) ^37^.

#### FinnGen

We downloaded time-aware (stemming from longitudinal cohort data) risk estimates from the FinnGen Risteys R10 portal for six neurodegenerative diseases: Alzheimer’s disease, amyotrophic lateral sclerosis, generalized dementia, multiple sclerosis, Parkinson’s disease, and vascular dementia. [FinnGen endpoints](https://www.finngen.fi/en/researchers/clinical-endpoints) are collections of one or more ICD10 codes, as defined by FinnGen clinical expert groups. In FinnGen all concurrent endpoints include at least 10 cases.

### ADDITIONAL STATISTICAL ANALYSES

#### Regression models

The Cox proportional hazard model was used to calculate HRs for Tables 2-4. Statsmodels GLM was used for Table 5. All code can be found at: [https://github.com/NIH-CARD/NDD_x_sleep].

#### Polygenic risk score calculation

Polygenic risk scores (PRSs) were calculated using summary statistics for AD (Kunkle et al) and PD (Nalls et al) ^31,32^. Summary statistics did not contain UK Biobank individuals. Interaction terms were defined as a binarized G47 or F51 code with one added to it to avoid scaling issues when multiplied by the PD or AD PRS (the PD and AD PRS were Z scaled with no true zero values at floating point limits).

